# Geospatial Analysis and Determinant Factors of Comorbidity Presence in Patients with Diabetes

**DOI:** 10.1101/2024.06.15.24308978

**Authors:** Víctor Juan Vera-Ponce, Fiorella E. Zuzunaga-Montoya, Luisa Erika Milagros Vásquez-Romero, Joan A. Loayza-Castro, Nataly Mayely Sanchez-Tamay, Carmen Inés Gutierrez De Carrillo

## Abstract

**Introduction:** The prevalence of diabetes mellitus (DM) has shown a significant increase in recent decades, leading to a rise in associated complications.

**Objective:** To explore the determinant factors and geographical distribution of comorbidities and their number in patients with diabetes in Peru.

**Methods:** Cross-sectional study based on a database providing detailed demographic and clinical information on DM patients affiliated with the Comprehensive Health Insurance (SIS: acronym in spanish) in Peru. The dependent variables in this study are twofold: the type of comorbidities present in DM patients and the number of comorbidities they have. Comorbidities were categorized into three groups: DM with obesity/dyslipidemia, DM with hypertension, and DM with mental health disorders. The number of comorbidities was classified as none, one, two, or three comorbidities.

**Results:** A total of 1,355,354 patients were included. Male patients, older individuals, and those with a longer time since diagnosis have different probabilities of presenting the comorbidities and a higher number of them. Additionally, the geospatial analysis showed apparent regional variations in the prevalence and number of comorbidities, highlighting the influence of environmental and socioeconomic factors and access to healthcare services.

**Conclusions:** This study identified significant demographic and clinical factors associated with comorbidities in patients with DM in Peru. These findings showed the need for personalized, region-specific diabetes management. Therefore, public health policies should adapt to meet the needs of different regions and groups. Improving healthcare access is crucial, especially where comorbidity prevalence is high. Further education programs must address diet and exercise comorbidities, focusing on vulnerable people.

## Introduction

Diabetes mellitus (DM) affects more than 463 million people worldwide; the number is projected to increase significantly to reach 700 million by the year 2045, according to the International Diabetes Federation ^(1)^. Globally, DM represents a leading cause of mortality and has been linked to comorbidities, including obesity, dyslipidemia, hypertension (HTN) ^(2)^, and mental health disorders ^(3)^. These multiple physical and psychosocial factors have also demonstrated significant negative impacts on patients’ quality of life while increasing healthcare system costs significantly ^(4,5)^.

Similarly, in Peru, during the last years, there was an essential rise in diabetes prevalence, which led to the increment of its associated complications, especially among low-socioeconomic population groups. In addition, the Peruvian context presents marked socioeconomic differences and significant disparities regarding access to healthcare services across different regions. Consequently, it is relevant to study and examine factors contributing to comorbidity ^(6)^.

As observed globally, the increasing rate of comorbidities is a significant concern. Understanding their presence and various determinant factors, as well as their distributions in different provinces, would be valuable in appreciating their impact on health outcomes. This understanding could also identify areas that need urgent interventions through the efficient allocation of resources by way of tailored prevention and treatment programs based on evidence from this study ^(7–9)^.

Therefore, this study aimed to combine statistical and geospatial analyses to provide a comprehensive view of the determinant factors and geographical distribution of comorbidities in DM patients affiliated with SIS in Peru. Additionally, we explored how comorbidities vary based on these factors and their spatial distribution across Peruvian provinces.

## Methods

### Type and Study Design

This is an analytical cross-sectional study based on an open-access database providing detailed demographic and clinical information on DM patients affiliated with the Comprehensive Health Insurance (SIS: acronym in spanish) in Peru.

### Population, Sample, and Eligibility Criteria

The study included patients affiliated with SIS with a confirmed diagnosis of DM (type 1, type 2, or other types of diabetes). It excluded those without data on the presence of any comorbidities.

### Variables and Measurement

The dependent variables in this study are twofold: the type of comorbidities present in DM patients and the number of comorbidities they have. Comorbidities were categorized into three groups: DM with obesity/dyslipidemia, DM with HTN, and DM with mental health disorders. The number of comorbidities was classified as none, one, two, or three comorbidities.

The potential determinant factors included the patient’s sex, categorized as male or female. The patient’s age was grouped into categories: under 18 years, then in 10-year intervals up to 70 years or older. The geographic region of residence was classified as Metropolitan Lima, the rest of the coast, highlands, and jungle. The time since diagnosis, I was referring to the duration since DM diagnosis was categorized in annual intervals of up to 6 years or more. Additionally, the type of diagnosed DM was grouped into type 1 DM (T1DM), type 2 DM (T2DM), and other specific types of diabetes.

### Procedures

This study accessed data from active SIS members in Peru diagnosed with diabetes mellitus from the country’s public open data repository ^(10)^. This dataset includes individual information and details about medical consultations received in the last quarter. The obtained data were cleaned, and pertinent variables for the analysis were selected.

Patients in the repository were categorized according to their type of DM (type 1, type 2, or others) and whether they exhibited HTN. Their glucose levels, lipids, and other laboratory variables were examined. Their body mass index (BMI) was determined to identify those with obesity (BMI ≥ 30 kg/m²), and their psychological well-being was evaluated. These assessments were conducted in the regions where SIS operates, and the results were documented in the repository used for this study.

The study sample comprised patients enrolled in the Comprehensive Health Insurance (SIS) with a confirmed diagnosis of DM who had received at least one medical consultation in the last quarter. Patients whose DM diagnosis did not specify the type (1, 2, or other) were excluded to ensure data accuracy. The International Classification of Diseases, Tenth Revision (ICD-10) was used to categorize comorbidities. DM with obesity/dyslipidemia was categorized under ICD-10 codes E660-E669 and E780-E785; DM with hypertension under codes I10X, I150, I151, I152, I158, and I159; and DM with mental health disorders under codes F000-F09X and F100-F259, F300-F99X, respectively.

### Statistical Analysis

Data analysis was performed using R software, version 4.3.1. First, a descriptive analysis of the demographic and clinical characteristics of the patients included in the study was conducted. Categorical variables were summarized using absolute and relative frequencies, expressed as percentages.

Subsequently, two multivariable analysis models were developed to identify the determinant factors of comorbidities in patients with diabetes. The first model focused on the type of comorbidities, evaluating the relationship between independent variables and each kind of comorbidity using Poisson regression with robust variance. Using multinomial logistic regression, the second model analyzed the same factors with the number of comorbidities (none versus one, two, or three).

Additionally, a geospatial analysis was conducted to evaluate the geographical distribution of comorbidities in the provinces of Peru. This analysis included several steps. First, the prevalence of each type of comorbidity and the number of comorbidities by province were calculated and then integrated with the shapefile of the regions of Peru. Heatmaps were then created using ggplot2 to visualize the prevalence distribution.

Both global and local Moran’s I indices were used to explore spatial autocorrelation at the provincial level, with a significance level of 0.05 using 999 permutations. These permutations generate a null distribution of spatial autocorrelation values under the hypothesis of no spatial clustering. This allows for calculating p-values and evaluating the statistical significance of observed spatial patterns. The correlation visualization was performed using a scatter plot with values ranging from −1 to +1, assessing whether the units of analysis tend toward clustering (positive autocorrelation), dispersion (negative autocorrelation), or randomization. Values greater than zero indicate spatial clustering, values equal to zero suggest random distribution and values less than zero indicate spatial dispersion.

The spatial representation was conducted using the local Moran’s I index (LISA), which identifies five types of clustering: 1) provincial clusters with high prevalence of comorbidities surrounded by provinces with above-average prevalence (“high-high” or hot spots); 2) provincial clusters with high prevalence surrounded by provinces with below-average prevalence (“high-low”); 3) provincial clusters with low prevalence surrounded by provinces with above-average prevalence (“low-high”); 4) clusters with low provincial prevalence of comorbidities surrounded by provinces with low prevalence (“low-low”); and 5) provinces with prevalence that does not significantly correlate with surrounding provinces (“non-significant”).

### Ethical Considerations

The study adhered to all current ethical and privacy regulations. Despite using an open-access database, patient confidentiality and anonymity were ensured. Therefore, ethical approval from a research ethics committee was not deemed necessary.

## Results

A total of 1,355,354 patients were included in the study. The majority of patients were female, representing 66.13% of the sample. In terms of age, the largest group comprised patients aged 50 to 59 years (24.35%). Regarding the type of DM, only 6.67% had type 1 DM. Geographically, most patients resided in Metropolitan Lima (37.37%). Concerning the time since the diabetes diagnosis, 21.31% of patients had been diagnosed between 1 and 2 years before the study. In terms of comorbidities, 64.46% of patients had diabetes along with obesity/dyslipidemia. Finally, considering the comorbidities, only 9.14% had all three comorbidities studied.

**Table 1.** Descriptive Characteristics of the Study Sample.

| Características | n = 1,355,354 |
| --- | --- |
| **Sex** |  |
| Female | 896,302 (66.13%) |
| Male | 459,052 (33.87%) |
| **Edad** |  |
| Under 18 years | 30,146 (2.22%) |
| 18 - 29 years | 63,267 (4.67%) |
| 30 - 39 years | 104,003 (7.67%) |
| 40 - 49 years | 218,914 (16.15%) |
| 50 - 59 years | 329,998 (24.35%) |
| 60 - 69 years | 321,288 (23.71%) |
| 70 years and older | 287,494 (21.22%) |
| **Type of Diabetes** |  |
| Type 1 diabetes | 90,380 (6.67%) |
| Type 2 diabetes | 1,244,198 (91.80%) |
| Other specified types of DM | 20,776 (1.53%) |
| **Region** |  |
| Metropolitan Lima | 506,526 (37.37%) |
| Rest of the Coast | 496,984 (36.67%) |
| Highlands | 189,034 (13.95%) |
| Jungle | 162,810 (12.01%) |
| **Time since diagnosis** |  |
| 0 to 1 year | 50,569 (3.73%) |
| 1 to 2 year | 288,794 (21.31%) |
| 2 to 3 year | 239,160 (17.65%) |
| 3 to 4 year | 149,786 (11.05%) |
| 4 to 5 year | 202,075 (14.91%) |
| 5 to 6 year | 243,926 (18.00%) |
| More than six years | 181,044 (13.36%) |
| **DM + Obesity/Dyslipidemia** |  |
| No | 481,688 (35.54%) |
| Yes | 873,666 (64.46%) |
| **DM + Hypertension** |  |
| No | 830,800 (61.30%) |
| Yes | 524,554 (38.70%) |
| **DM + Mental Health Disorder** |  |
| No | 1,060,093 (78.22%) |
| Yes | 295,261 (21.78%) |
| **Number of Comorbidities** |  |
| None | 300,438 (22.17%) |
| One | 540,215 (39.86%) |
| Two | 390,837 (28.84%) |
| Three | 123,864 (9.14%) |

### Multivariable Analysis of Factors Associated with Each Type of Comorbidity

For the presence of obesity/dyslipidemia, male patients had a lower probability of having diabetes along with obesity/dyslipidemia compared to female patients. Regarding age, as age increased, so did the probability of presenting this comorbidity, with the highest probability observed in the 50 to 69 age groups. Patients with type 2 diabetes had a higher probability of having obesity/dyslipidemia compared to those with type 1 diabetes. Regarding region, patients from the Jungle showed a lower probability of having this comorbidity compared to those from Metropolitan Lima—a more extended time since diagnosis was associated with an increased probability of presenting obesity/dyslipidemia. Patients with hypertension and mental health disorders also showed a higher probability of having obesity/dyslipidemia.

Male patients had a similar probability of presenting hypertension compared to female patients. The probability of having diabetes along with hypertension increased notably with age, being incredibly high in the older age groups (60 years and above). Patients with type 2 diabetes and other specified types of diabetes had a lower probability of presenting hypertension compared to patients with type 1 diabetes. Patients from the Jungle showed a higher probability of having this comorbidity than those from Metropolitan Lima. A longer time since diagnosis was also associated with a higher probability of having hypertension. Patients with mental health disorders and obesity/dyslipidemia had a higher probability of presenting hypertension.

Regarding mental health disorders, male patients had a lower probability of presenting this comorbidity compared to female patients. The probability of having diabetes along with mental health disorders decreases with age. Patients with type 2 diabetes and other specified types of diabetes showed a lower probability of having mental health disorders compared to patients with type 1 diabetes. Patients from the Jungle also showed a lower probability of presenting this comorbidity than those from Metropolitan Lima—a more extended time since diagnosis was associated with an increased probability of delivering mental health disorders. Patients with hypertension and obesity/dyslipidemia also had a higher probability of having mental health disorders.

**Table 2.** Multivariate analysis of factors associated with comorbidities.

| Characteristic | Obesity/Dyslipidemia |  | Hypertension |  | Mental Health Disorder |  |
| --- | --- | --- | --- | --- | --- | --- |
|  | PRa | 95% CI | PRa | 95% CI | PRa | 95% CI |
| **Sex** |  |  |  |  |  |  |
| Female | Ref. |  | Ref. |  | Ref. |  |
| Male | 0.81 | 0.80, 0.81 | 0.98 | 0.97, 0.98 | 0.75 | 0.75, 0.76 |
| **Edad** |  |  |  |  |  |  |
| Under 18 years | Ref. |  | Ref. |  | Ref. |  |
| 18 - 29 years | 1.2 | 1.18, 1.22 | 3.1 | 2.87, 3.34 | 0.87 | 0.85, 0.89 |
| 30 - 39 years | 1.38 | 1.36, 1.40 | 4.81 | 4.47, 5.17 | 0.72 | 0.71, 0.74 |
| 40 - 49 years | 1.51 | 1.49, 1.53 | 7.98 | 7.44, 8.57 | 0.67 | 0.65, 0.68 |
| 50 - 59 years | 1.55 | 1.53, 1.57 | 12.2 | 11.4, 13.1 | 0.63 | 0.62, 0.64 |
| 60 - 69 years | 1.55 | 1.53, 1.57 | 17.4 | 16.2, 18.7 | 0.61 | 0.60, 0.62 |
| 70 years and older | 1.46 | 1.44, 1.48 | 23.5 | 21.9, 25.2 | 0.67 | 0.65, 0.68 |
| **Type of Diabetes** |  |  |  |  |  |  |
| Type 1 diabetes | Ref. |  | Ref. |  | Ref. |  |
| Type 2 diabetes | 1.2 | 1.19, 1.21 | 0.85 | 0.85, 0.86 | 0.87 | 0.86, 0.88 |
| Other specified types of DM | 1.14 | 1.13, 1.16 | 0.87 | 0.85, 0.88 | 0.91 | 0.89, 0.94 |
| **Region** |  |  |  |  |  |  |
| Metropolitan Lima | Ref. |  | Ref. |  | Ref. |  |
| Rest of the Coast | 1.03 | 1.03, 1.04 | 1.1 | 1.09, 1.10 | 0.76 | 0.76, 0.77 |
| Highlands | 1.02 | 1.01, 1.02 | 0.9 | 0.90, 0.91 | 0.77 | 0.77, 0.78 |
| Jungle | 0.82 | 0.82, 0.83 | 1.23 | 1.22, 1.23 | 0.71 | 0.70, 0.72 |
| **Time since diagnosis** |  |  |  |  |  |  |
| 0 to 1 year | Ref. |  | Ref. |  | Ref. |  |
| 1 to 2 year | 1.14 | 1.13, 1.15 | 1.07 | 1.05, 1.08 | 1.20 | 1.18, 1.23 |
| 2 to 3 year | 1.19 | 1.17, 1.20 | 1.15 | 1.13, 1.16 | 1.29 | 1.26, 1.32 |
| 3 to 4 year | 1.12 | 1.11, 1.13 | 1.2 | 1.19, 1.22 | 1.31 | 1.28, 1.34 |
| 4 to 5 year | 1.27 | 1.26, 1.28 | 1.12 | 1.11, 1.14 | 1.35 | 1.32, 1.38 |
| 5 to 6 year | 1.31 | 1.30, 1.32 | 1.15 | 1.13, 1.16 | 1.48 | 1.45, 1.52 |
| More than six years | 1.39 | 1.38, 1.41 | 1.28 | 1.27, 1.30 | 1.58 | 1.54, 1.61 |
| **DM + Hypertension** |  |  |  |  |  |  |
| No | Ref. |  | — | — | Ref. |  |
| Yes | 1.18 | 1.18, 1.18 | — | — | 1.56 | 1.55, 1.57 |
| **DM + Mental Health Disorder** |  |  |  |  |  |  |
| No | Ref. |  | Ref. |  | — | — |
| Yes | 1.17 | 1.17, 1.17 | 1.3 | 1.30, 1.31 | — | — |
| **DM + Obesity/Dyslipidemia** |  |  |  |  |  |  |
| No | — | — | Ref. |  | Ref. |  |
| Yes | — | — | 1.34 | 1.33, 1.35 | 1.58 | 1.57, 1.59 |

\*The other variables were independently adjusted for each factor.
aRP: Adjusted Prevalence ratio. 95% CI: 95% confidence interval
Source: self-made

### Multivariable Analysis of Factors Associated with the Number of Comorbidities

Male patients had a lower probability of presenting one comorbidity than female patients. As age increased, so did the probability of delivering this, with the highest probability observed in the 50 to 69 age groups. Patients with type 2 diabetes and other specified types of diabetes had a higher probability of having one comorbidity compared to patients with type 1 diabetes. Regarding region, patients from the rest of the Coast showed a higher probability of having one comorbidity than those from Metropolitan Lima. In comparison, patients from the Highlands and Jungle showed a lower probability—a more extended time since diagnosis was associated with an increased probability of presenting one comorbidity.

In the case of those presenting two comorbidities, male patients also had a lower probability of explaining this than female patients. The probability of having two comorbidities increased significantly with age, especially in the older age groups (60 years and above). Patients with type 2 diabetes and other specified types of diabetes had a higher probability of presenting two comorbidities compared to patients with type 1 diabetes. Patients from the rest of the Coast showed a higher probability of having two comorbidities than those from Metropolitan Lima. In comparison, patients from the Highlands and Jungle showed a lower probability. A longer time since diagnosis was also associated with a higher probability of having two comorbidities.

Regarding three comorbidities, male patients had a lower probability of presenting this condition than female patients. The probability of having three comorbidities increased notably with age, being extremely high in the oldest age groups (70 years and above). Patients with type 2 diabetes and other specified types of diabetes did not show significant differences in the probability of presenting three comorbidities compared to patients with type 1 diabetes. Patients from the rest of the Coast showed a lower probability of having three comorbidities than those from Metropolitan Lima. In comparison, patients from the Highlands and Jungle showed an even lower probability—a more extended time since diagnosis was significantly associated with an increased likelihood of presenting three comorbidities.

**Table 3.**
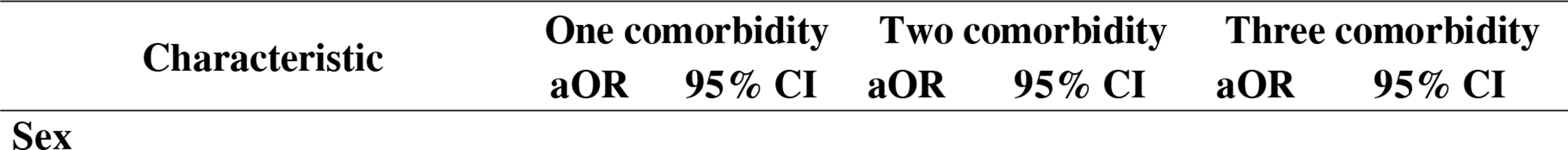
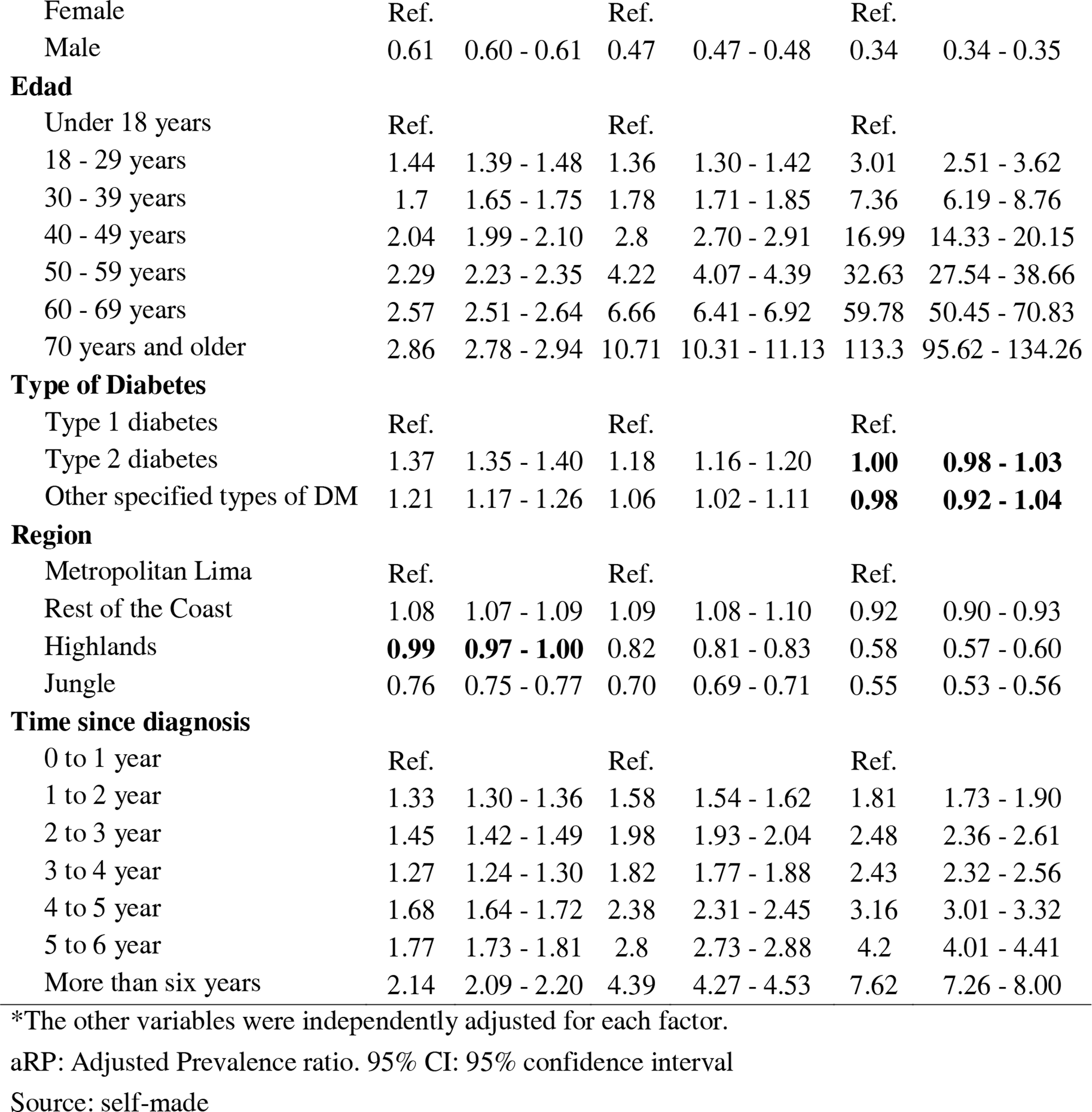
Multivariable Analysis of Factors Associated with the Number of Comorbidities.

| Characteristic | One comorbidity |  | Two comorbidity |  | Three comorbidity |  |
| --- | --- | --- | --- | --- | --- | --- |
|  | aOR | 95% CI | aOR | 95% CI | aOR | 95% CI |
| Sex |  |  |  |  |  |  |
| Female | Ref. |  | Ref. |  | Ref. |  |
| Male | 0.61 | 0.60 - 0.61 | 0.47 | 0.47 - 0.48 | 0.34 | 0.34 - 0.35 |
| **Edad** |  |  |  |  |  |  |
| Under 18 years | Ref. |  | Ref. |  | Ref. |  |
| 18 - 29 years | 1.44 | 1.39 - 1.48 | 1.36 | 1.30 - 1.42 | 3.01 | 2.51 - 3.62 |
| 30 - 39 years | 1.7 | 1.65 - 1.75 | 1.78 | 1.71 - 1.85 | 7.36 | 6.19 - 8.76 |
| 40 - 49 years | 2.04 | 1.99 - 2.10 | 2.8 | 2.70 - 2.91 | 16.99 | 14.33 - 20.15 |
| 50 - 59 years | 2.29 | 2.23 - 2.35 | 4.22 | 4.07 - 4.39 | 32.63 | 27.54 - 38.66 |
| 60 - 69 years | 2.57 | 2.51 - 2.64 | 6.66 | 6.41 - 6.92 | 59.78 | 50.45 - 70.83 |
| 70 years and older | 2.86 | 2.78 - 2.94 | 10.71 | 10.31 - 11.13 | 113.3 | 95.62 - 134.26 |
| **Type of Diabetes** |  |  |  |  |  |  |
| Type 1 diabetes | Ref. |  | Ref. |  | Ref. |  |
| Type 2 diabetes | 1.37 | 1.35 - 1.40 | 1.18 | 1.16 - 1.20 | **1.00** | **0.98 - 1.03** |
| Other specified types of DM | 1.21 | 1.17 - 1.26 | 1.06 | 1.02 - 1.11 | **0.98** | **0.92 - 1.04** |
| **Region** |  |  |  |  |  |  |
| Metropolitan Lima | Ref. |  | Ref. |  | Ref. |  |
| Rest of the Coast | 1.08 | 1.07 - 1.09 | 1.09 | 1.08 - 1.10 | 0.92 | 0.90 - 0.93 |
| Highlands | **0.99** | **0.97 - 1.00** | 0.82 | 0.81 - 0.83 | 0.58 | 0.57 - 0.60 |
| Jungle | 0.76 | 0.75 - 0.77 | 0.70 | 0.69 - 0.71 | 0.55 | 0.53 - 0.56 |
| **Time since diagnosis** |  |  |  |  |  |  |
| 0 to 1 year | Ref. |  | Ref. |  | Ref. |  |
| 1 to 2 year | 1.33 | 1.30 - 1.36 | 1.58 | 1.54 - 1.62 | 1.81 | 1.73 - 1.90 |
| 2 to 3 year | 1.45 | 1.42 - 1.49 | 1.98 | 1.93 - 2.04 | 2.48 | 2.36 - 2.61 |
| 3 to 4 year | 1.27 | 1.24 - 1.30 | 1.82 | 1.77 - 1.88 | 2.43 | 2.32 - 2.56 |
| 4 to 5 year | 1.68 | 1.64 - 1.72 | 2.38 | 2.31 - 2.45 | 3.16 | 3.01 - 3.32 |
| 5 to 6 year | 1.77 | 1.73 - 1.81 | 2.8 | 2.73 - 2.88 | 4.2 | 4.01 - 4.41 |
| More than six years | 2.14 | 2.09 - 2.20 | 4.39 | 4.27 - 4.53 | 7.62 | 7.26 - 8.00 |

\*The other variables were independently adjusted for each factor.
aRP: Adjusted Prevalence ratio. 95% CI: 95% confidence interval
Source: self-made

**Figure 1** shows the heat maps of the prevalence of patients with diabetes mellitus (DM) in combination with three comorbidities: obesity/dyslipidemia (A), hypertension (HTN) (B), and mental health disorders (C). In map A, darker green areas represent a higher prevalence of obesity/dyslipidemia, highlighting the central and southern coastal regions. Map B uses shades of orange to show the prevalence of HTN, with the highest prevalence in the northern and some central areas. Map C employs purple tones, indicating higher prevalence in certain southern provinces.

**Figure 2** presents the spatial representation maps using LISA analysis for patients with DM along with comorbidities of obesity/dyslipidemia (A), HTN (B), and mental health disorders (C). These maps display the significant spatial clusters of high and low prevalence for each comorbidity in the provinces. Moran’s calculated index in map A was 0.3621 (p<0.001), indicating a strong positive spatial autocorrelation. In map B, Moran’s index was 0.3742 (p<0.001), indicating a strong positive spatial autocorrelation. Finally, Moran’s index in map C was 0.3095 (p<0.001), suggesting significant positive spatial autocorrelation.

**Figure 3** shows the heat maps of the prevalence of patients with DM and different numbers of comorbidities: none (A), one (B), two (C), and three comorbidities (D). In map A, orange tones show the prevalence of patients without comorbidities, with the highest prevalence observed in some southern provinces. In green, Map B indicates the regions where DM patients have one comorbidity, highlighting central and south coast areas. In map C, purple tones show the prevalence of patients with two comorbidities, with a high concentration in the north and some places in the south. Finally, in map D, blue tones represent the prevalence of patients with three comorbidities, showing higher prevalence in the Amazon region.

**Figure 4** presents the spatial representation maps using LISA analysis for patients with diabetes mellitus (DM) with none (A), one (B), two (C), and three comorbidities (D). Moran’s calculated index for map A was 0.2965 (p<0.001), indicating a strong positive spatial autocorrelation. The Moran’s index for map B was 0.2171 (p<0.001), indicating positive spatial autocorrelation. For map C, Moran’s index was 0.3045 (p<0.001), suggesting considerable positive spatial autocorrelation. The maps show high and low prevalence clusters, with areas in red and blue, respectively, identifying regions with two comorbidities. Finally, the Moran’s index for map D was 0.2733 (p<0.001), indicating positive spatial autocorrelation. The maps in blue and red tones show areas with high and low prevalence of three comorbidities.

## Discussion

### Prevalence of Types and Number of Comorbidities

The results revealed a significant proportion of the entire population examined to have DM along with additional comorbidities that were more common and frequently occurred compared to other medical conditions in some cases. Specifically, the prevalence of patients with DM simultaneously presenting obesity/dyslipidemia was relatively high (64.46%). These findings agree with observations made by other investigators ^(2,11–13)^.; however, several studies noted HTN affecting more diabetics as opposed to DM coexisting on its own or together with other underlying disease processes ^(7,14)^. Possible reasons for this variability may be influenced by genetic, environmental, and socioeconomic factors, as well as variations in healthcare systems and treatment practices. The higher prevalence of obesity/dyslipidemia indicates the strong relationship between diabetes and metabolic alterations, often sharing common risk factors such as unhealthy diets and lack of physical exercise. The literature has found that the coexistence of obesity/dyslipidemia with DM complicates diabetes management, increasing the risk of cardiovascular complications and worsening the overall prognosis of patients ^(15,16)^.

HTN was also a frequent comorbidity, affecting 38.70% of DM patients. Therefore, people who develop DM simultaneously with HTN lose their chance to prevent CVD complications and are at higher probability for kidney failure, CVD including MI, heart stroke, and other severe cardiovascular complications than people suffering from single conditions. Due to this, rigorous blood pressure control in diabetics should also be practiced thoroughly, focusing specifically on healthy lifestyle education along with proper use of antihypertensive drugs being used by diabetic patients ^(2,17)^.

On the other hand, mental health disorders were less prevalent, with 21.78% of DM patients reporting this comorbidity. Although this figure is lower than the other comorbidities studied, it remains significant ^(18,19)^. Mental health disorders such as depression and anxiety can complicate diabetes management by interfering with treatment adherence and motivation for self-care. DM patients with mental health issues require an integrated treatment approach that addresses both the physical and psychological aspects of their health ^(20)^.

Regarding the number of comorbidities, 39.86% of DM patients had one comorbidity, 28.84% had two comorbidities, and 9.14% had three comorbidities. This indicates that a significant proportion of DM patients face an additional burden of chronic conditions, further complicating their management and treatment ^(21)^. Although the number of patients with three comorbidities is still below 10%, it is higher than other studies, which could indicate an increase in these as the years go by ^(13)^. The presence of multiple comorbidities not only increases the risk of severe complications but also requires a more complex and multidisciplinary approach to care ^(22)^. These patients may experience a poorer quality of life and higher healthcare costs. Therefore, health systems must implement comprehensive and coordinated management strategies that address all chronic conditions in these patients, ensuring continuous and personalized care that can improve health outcomes and reduce the disease burden.

### Sex and Comorbidities

In our study, male patients were found to be less likely to present any of the studied comorbidities, such as obesity/dyslipidemia, hypertension, and mental health disorders, compared to female patients. Additionally, men also showed a lower probability of having multiple comorbidities. Several pathophysiological and behavioral reasons can explain this finding. From a pathophysiological perspective, body fat distribution and sex hormones play an essential role. Men tend to accumulate more visceral fat, which is more associated with increased metabolic risk but can also be more easily mobilized during weight loss or increased physical activity. On the other hand, estrogens in premenopausal women have protective effects against cardiovascular and metabolic diseases. Still, after menopause, the decline in estrogen levels increases the risk of developing these comorbidities ^(23,24)^.

In terms of behavior and lifestyle, women tend to use healthcare services more frequently than men, which can lead to earlier and more frequent diagnoses of comorbidities. This higher utilization of healthcare services could result in a higher registration of comorbidities in women. Additionally, women tend to be more attentive to their health and follow medical recommendations more rigorously than men ^(25)^. Men, on the other hand, may underestimate symptoms and not seek medical attention until more advanced stages of the disease, which could lead to lower detection of comorbidities compared to women ^(23)^.

Finally, psychosocial factors can also contribute to these differences. Women tend to report higher levels of stress and mental health issues, which can contribute to the occurrence of other comorbidities. Social and cultural roles also influence how men and women manage their health. Women may be more willing to seek help and support, while men might feel pressured to maintain an image of strength and not seek medical assistance ^(3,23)^.

### Age and Comorbidities

Our study found a significant relationship between age and the probability of presenting comorbidities in DM patients. This means that as we move up one unit in each additional age category, patients with diabetes mellitus have a higher probability of showing obesity/dyslipidemia, as well as other hypertension-related issues. Specifically, patients are at a greater risk for developing hypertension as they age, with a strong correlation coefficient in older adults over 65 years old due to the progressive process involving arterial stiffness as a significant contributing factor ^(26)^. Additionally, visceral fat accumulates steadily with age, altering the entire lipid metabolism and favoring even more severe cardiovascular diseases such as atherosclerosis. Other physiological changes regarding the cardiovascular system may be minor but seem less significant in this context ^(27)^.

In contrast, older patients were found to be less likely to present mental health disorders compared to younger patients. This finding could be related to several factors. First, mental health disorders may be more commonly diagnosed and reported in early life stages due to a greater focus on emotional and psychological development during childhood and adolescence. Additionally, older adults may be less likely to seek help or receive a diagnosis for mental health issues due to generational stigmas and differences in the perception of mental health. Lastly, physical comorbidities may receive more attention in older adults, overshadowing the diagnosis of mental conditions ^(28,29)^.

Furthermore, it was found that older age is associated with a higher probability of having more comorbidities. This increase may be due to the accumulation of risks over a lifetime and the physiological deterioration that occurs with aging. Over time, prolonged exposure to risk factors such as physical inactivity, inadequate diet, and chronic stress can contribute to the simultaneous development of multiple comorbidities. Additionally, the body’s decreased ability to maintain homeostasis and repair cellular damage with age can lead to deterioration in various organ systems, facilitating the concurrent appearance of several chronic conditions ^(28,30,31)^.

### Type of Diabetes and Comorbidities

Our study found that patients with T2DM and other specific types of diabetes are more likely to present obesity/dyslipidemia but less likely to present HTN compared to patients with T1DM. This difference could be due to several pathophysiological and disease management factors. Patients with T2DM often present a phenotype of insulin resistance associated with central obesity and dyslipidemia, less common in T1DM, where absolute insulin deficiency predominates ^(32,33)^. The lower probability of HTN in T2DM and other specific types of diabetes could be related to the higher prevalence of HTN in T1DM due to the coexistence of autoimmune factors and possibly greater arterial stiffness from an earlier age in these patients ^(32,34)^.

Additionally, it was found that patients with T2DM and other specific types of diabetes are less likely to present mental health disorders compared to patients with T1DM. This difference may be attributed to several factors. Patients with T1DM are generally diagnosed at younger ages, implying a prolonged and potentially more severe impact on emotional and mental well-being due to the need for constant and rigorous disease management from a young age. Additionally, the burden of living with a chronic autoimmune disease can increase vulnerability to mental health problems. In contrast, patients with T2DM are usually diagnosed in adulthood and may have more developed coping mechanisms and a different perception of their health condition ^(23)^.

Regarding the number of comorbidities, it was found that patients with T2DM and other specific types of diabetes are more likely to have one or two comorbidities than patients with T1DM. However, these groups had no significant differences in the probability of presenting three comorbidities. This trend can be explained by the progressive and multisystemic nature of T2DM, which is often associated with a range of metabolic and cardiovascular complications due to insulin resistance and obesity ^(22)^. On the other hand, the absence of significant differences in the presence of three comorbidities suggests that once a certain threshold of multisystemic deterioration is reached, the differences between types of diabetes may become less pronounced. This underscores the need for comprehensive and personalized management of comorbidities in all diabetic patients, regardless of type ^(9)^.

### Time Since Diagnosis and Comorbidities

Our study found that the longer the time since the diabetes diagnosis, the more likely patients are to present any studied comorbidities. Additionally, as the time since diagnosis increased, so did the probability of having more comorbidities. This finding can be explained by several pathophysiological reasons related to disease progression. Over time, DM tends to progress, and prolonged accumulation of chronic hyperglycemia can lead to multisystem damage. Continuous exposure to high blood glucose levels can damage blood vessels and nerves, contributing to the development of complications such as HTN and mental health disorders. Additionally, prolonged hyperglycemia can induce metabolic changes that increase the risk of obesity and dyslipidemia ^(21,22)^.

Moreover, patients’ ability to maintain homeostasis and repair cellular damage decreases as they age. Aging itself is a risk factor for many of the comorbidities associated with diabetes. Decreased renal function, increased arterial stiffness, and reduced muscle mass and cognitive function with age can contribute to the development of HTN, obesity/dyslipidemia, and mental health disorders ^(27)^.

Time since diagnosis can also influence health behaviors and treatment adherence. Over time, patients may experience “treatment fatigue,” leading to lower adherence to medication regimens and less rigorous diet and exercise control. This decline in active diabetes management can increase the likelihood of developing comorbidities. Additionally, the psychological burden of living with a chronic disease for an extended period can contribute to mental health problems, which in turn can negatively impact the management of diabetes and other comorbidities ^(35,36)^.

### Region and Comorbidities

Our study found that the region type significantly influences the probability of presenting comorbidities in DM patients. Patients residing in the coast and jungle regions are more likely to present HTN than patients from Metropolitan Lima, while patients from the highlands are less likely to present HTN. This finding can be explained by lifestyle and cardiovascular health differences between different areas, specifically urban and rural zones ^(37)^. Additionally, diets in these regions may include high salt and processed foods, increasing the risk of HTN ^(38)^. In contrast, the altitude of the highlands induces cardiovascular adaptations that potentially protect against HTN, and physical activity is often more frequent due to the demands of daily life in these areas ^(39)^.

Additionally, it was found that patients from all regions (coast, highlands, and jungle) are less likely to present mental health disorders compared to those from Metropolitan Lima. This difference may be related to socioeconomic factors and access to healthcare services. Metropolitan Lima has more accessible and more frequent access to mental health services, which can lead to a higher diagnosis of mental health disorders. Additionally, urbanization and the stress associated with living in a large city contribute to a higher prevalence of mental health problems ^(40)^. In contrast, in the coastal, highland, and jungle regions, there may be fewer diagnoses of mental health disorders due to lower access to specialized health services and possibly less demand or awareness of mental health ^(7)^.

Finally, it was observed that patients from the coast, highlands, and jungle regions are less likely to have multiple comorbidities than those from Metropolitan Lima. This finding may be due to differences in lifestyle, health perception, and access to healthcare services. In Lima, access to diagnoses and treatments is more advanced, which could result in higher detection of multiple comorbidities ^(41)^. Additionally, urban life may contribute to less healthy lifestyles, with less physical activity and poorer diets ^(37)^. On the other hand, rural regions may have a more significant stigma associated with mental health and less seeking of help, which can lead to lower rates of diagnosis and reporting of comorbidities.

### Geospatial Analysis of Types of Comorbidities

The geospatial analysis reveals that areas with a high prevalence of obesity/dyslipidemia in DM patients are predominantly concentrated in the Amazon region and the north of the country. These concentrations suggest that specific regional factors, such as diet, physical activity, and access to healthcare services, play a crucial role in the prevalence of obesity and dyslipidemia. These regions’ socioeconomic and cultural characteristics can also influence obesity and dyslipidemia patterns. On the other hand, areas of low prevalence are dispersed across various regions, which may indicate better health practices or access to preventive treatments in those areas ^(37)^.

The prevalence of HTN in DM patients shows a high concentration in the northern region and some areas of the central part of the country. These results suggest that hypertension in DM patients is influenced by geographic and environmental factors, such as a high-salt diet and a sedentary lifestyle, which are more common in coastal and jungle regions ^(42)^. The dispersion of low-prevalence areas may indicate that these regions have better health practices or access to preventive treatments. This underscores the need for specific interventions in areas with a high prevalence of HTN to address these risk factors.

Mental health disorders show a higher prevalence in some southern regions and the Amazon region. This suggests that regional factors, including access to mental health services, socioeconomic and cultural factors, and the level of urbanization influence mental health disorders. Concentrations in the south and Amazon regions may reflect a lower availability of mental health resources and a higher burden of stress associated with living conditions in these areas. Low-prevalence areas of mental health disorders are more dispersed, which could indicate better socioeconomic conditions and greater access to mental health services in those regions ^(40)^.

High prevalence areas indicate regions where DM patients are at greater risk of developing comorbidities, possibly due to specific regional factors such as diet, lifestyle, and access to healthcare. Low prevalence areas may reflect regions with better public health practices, greater access to preventive and treatment services, and better socioeconomic conditions ^(42)^. Identifying these patterns is crucial for planning specific public health interventions for regions with the greatest need for resources and prevention and treatment programs.

### Geospatial Analysis of the Number of Comorbidities

The geospatial analysis reveals that areas with a lower prevalence of comorbidities in DM patients are dispersed across several regions of the country. Low prevalence areas are mainly concentrated in the highlands and some parts of the jungle. This pattern may suggest that geographic and environmental factors, such as altitude and an active lifestyle, protect against multiple comorbidities ^(37)^. Additionally, health practices and access to medical services in these regions could contribute to a lower prevalence of comorbidities ^(41)^.

A higher prevalence of patients with one or more comorbidities is observed in coastal and jungle regions. This pattern suggests that lifestyle and diet-related factors in these regions may contribute to developing an additional comorbidity in DM patients. A high-salt and fat diet and lower physical activity levels may influence the concentration in these areas. Additionally, access and quality of healthcare services in these regions may affect the prevalence of comorbidities ^(43)^.

Furthermore, for patients with three comorbidities, the LISA analysis shows clusters of high prevalence in various regions, including the central and southern coast and some areas of the jungle. Three comorbidities in these patients indicate significant health deterioration, influenced by genetic, environmental, and behavioral factors ^(13,42,43)^.

### Limitations of the Study

This study presents several limitations that should be considered when interpreting the results. First, the study’s cross-sectional nature prevents establishing causality between the studied variables and the prevalence of comorbidities in DM patients. Second, unmeasured confounding factors may influence the observed association between demographic and clinical characteristics and the prevalence of comorbidities. Third, secondary databases, while valuable for obtaining large samples, can introduce biases due to the lack of specific and detailed information on important variables that were not recorded in the original database.

### Conclusions

This study identified significant demographic and clinical factors associated with comorbidities in DM patients in Peru. These findings showed the need for personalized, region-specific diabetes management. Therefore, public health policies should adapted to meet the needs of different regions and groups. Improving healthcare access is crucial, especially where comorbidity prevalence is high. Further education programs must address diet and exercise comorbidities, focusing on vulnerable people. More so, adapting clinical interventions will address region-specific needs and improve care for everyone affected by diabetes, yet challenges remain. Improving quality healthcare services is crucial; nevertheless, it is still difficult without the proper support from authorities.

## Acknowledgments

A special thanks to the members of the Tropical Diseases Research Institute, Universidad Nacional Toribio Rodríguez de Mendoza de Amazonas (UNTRM), Amazonas, Peru, for their support and contributions throughout the completion of this research.

## Informed consent

It was not necessary to obtain informed consent for this study.

## Data availability

The data supporting the findings of this study can be accessed by the original research paper at the follow link: https://www.datosabiertos.gob.pe/dataset/afiliados-activos-en-el-seguro-integral-de-salud-con-diagn%C3%B3stico-de-diabetes-mellitus-sis

## Authors’ contribution

Víctor Juan Vera-Ponce: Conceptualization, Investigation, Methodology, Resources, Writing - Original Draft, Writing - Review & Editing

Fiorella E. Zuzunaga-Montoya: Investigation, Project administration, Writing - Original Draft, Writing - Review & Editing

Nataly Mayely Sanchez-Tama: Investigation, Resources, Writing - Original Draft, Writing - Review & Editing

Joan A. Loayza-Castro: Software, Data Curation, Formal analysis, Writing - Review & Editing

Luisa Erika Milagros Vásquez-Romero: Validation, Visualization, Writing - Original Draft, Writing - Review & Editing

Carmen Inés Gutierrez De Carrillo: Methodology, Supervision, Funding acquisition, Writing - Review & Editing

